# Perioperative Dynamics of Autoantibodies Against Neurotransmitter Receptors in Liver Surgery: A Secondary Analysis of the PHYDELIO Trial

**DOI:** 10.1101/2025.07.10.25331255

**Authors:** A. Müller, C. Spies, H. Prüss, C. von Haefen, H. Heidecke, N. Paeschke, O. Wegwarth

**Author notes:** Correspondence: Prof. Dr. A. Müller, Charité Universitätsmedizin Berlin, Charitéplatz 1, 10117 Berlin.

## Abstract

**Background:** Postoperative delirium (POD) is a common neurocognitive complication following major surgery, particularly in older adults and those undergoing liver resections. Neuroinflammatory mechanisms are considered central to its pathophysiology, yet molecular mediators remain poorly defined. Autoantibodies (aABs) targeting G protein-coupled receptors (GPCRs)—especially those relevant to neurotransmission—may contribute to POD by disrupting neuroimmune homeostasis. This study explored the perioperative dynamics of GPCR-specific aABs and their association with POD incidence.

**Methods:** In this secondary analysis of the PHYDELIO randomized controlled trial (ISRCTN18978802), we evaluated serum aAB levels targeting five GPCRs (M3R, M4R, **β**_2_AR, D2R, and 5-HT_2_AR) in 142 patients undergoing liver surgery. Samples were collected preoperatively and on postoperative days 1, 2, and 7. POD was diagnosed using a comprehensive clinical assessment integrating validated screening tools and chart reviews. Repeated-measures ANOVAs examined time × group interactions, with additional post hoc and nonparametric tests applied as appropriate.

**Results:** Serum levels of four GPCR aABs (M3R, M4R, D2R, and 5-HT_2_AR) declined significantly following surgery and returned near baseline by day 7. **β**_2_AR aAB levels remained stable. Patients who developed POD (45.8%) exhibited consistently lower aAB levels, reaching statistical significance for M3R (p = .029). No significant time × delirium interaction was found for any antibody.

**Discussion:** Major abdominal surgery transiently alters GPCR aAB levels, suggesting perioperative immune modulation or adsorption to tissue following disrupted barriers. Lower M3R aAB concentrations were associated with POD, aligning with proposed cholinergic involvement in its pathogenesis. While only M3R-specific effects reached significance, the consistent trend across aABs supports further investigation into their role as biomarkers or mediators of POD. Future studies should assess their functional activity and potential utility in risk stratification and therapeutic targeting.

## INTRODUCTION

Postoperative delirium (POD) is a serious neurocognitive complication following major surgery, particularly in older adults and vulnerable patients undergoing extensive abdominal procedures such as liver resections. Clinically, POD is characterized by an acute and fluctuating disturbance in attention, cognition, and consciousness. It is associated with increased mortality, prolonged hospitalization, long-term cognitive decline, and a loss of independence.[1] POD can be understood as a form of perioperative single-organ failure affecting the brain, with an incidence ranging from approximately 6% to 20% following major surgery.[2] Despite growing clinical recognition and several proposed hypotheses — including inflammatory, neurotransmitter, and stress-related mechanisms — the underlying molecular pathogenesis of POD remains unclear. As a result, no causal treatment is currently available.

One factor that is increasingly recognized as a potential key contributor to the pathophysiology of postoperative delirium are neuroinflammatory mechanisms.[3] Surgical trauma and anesthetic exposure can trigger a systemic inflammatory response, which may compromise the integrity of the blood–brain barrier, disrupt neurotransmitter homeostasis, and activate resident immune cells within the central nervous system.[4] This pro-inflammatory cascade is thought to impair synaptic signaling and neural network connectivity—particularly in cholinergic, dopaminergic, and serotonergic pathways—contributing to the acute cognitive disturbances characteristic of POD.[5] Nevertheless, the specific immune mediators involved, and the factors underlying individual susceptibility, are not yet well understood.

One underexplored component of the neuroimmune interface in postoperative delirium involves functional autoantibodies (aABs) directed against G protein-coupled receptors (GPCRs) — the largest and most diverse family of membrane receptors in the human genome, encoded by over 800 genes. GPCRs are widely expressed not only on neurons but also on cells of the innate and adaptive immune systems, and they play a central role in maintaining physiological homeostasis across numerous organ systems. Notably, approximately one-third of all approved pharmaceuticals target GPCRs, underscoring their broad biological relevance.[6, 7]

From systemic autoimmune diseases and cardiovascular conditions to neuropsychiatric syndromes and post-viral sequelae, these antibodies have demonstrated their ability to function as predictors, mediators, and even potential therapeutic targets. It is hypothesized that autoantibodies targeting GPCRs disrupt intracellular signaling pathways, thereby impairing synaptic plasticity, neurotransmission, and neuroimmune homeostasis. This promotes neuroinflammation and contributes to the development of cognitive dysfunction.[8]

Autoantibodies targeting GPCRs are recognized as natural components of immunoregulation. However, under dysregulated conditions, these aABs may contribute to pathophysiological processes by acting either as agonists or antagonists, depending on their binding sites. For example, **β**_2_-adrenergic receptor (**β**_2_AR)-specific antibodies can exert inhibitory effects in allergic asthma or agonistic effects in disorders such as Chagas cardiomyopathy and Alzheimer’s disease—highlighting their context-dependent functional roles.[9, 10]

Importantly, GPCR-targeting aABs directed against receptors relevant to central nervous system function, including muscarinic acetylcholine receptors (M3R, M4R), dopamine D2 receptor (D2R), serotonin 2A receptor (5-HT2AR), and **β**_2_AR, have been implicated in conditions characterized by cognitive impairment, autonomic dysfunction, and neuroinflammation —such as chronic fatigue syndrome (CFS), neurodegenerative disorders, and autoimmune encephalopathies. [11] These findings suggest that GPCR autoantibodies may not only serve as biomarkers, but also as active mediators in the neurocognitive and inflammatory processes underlying disorders such as POD.

In this secondary analysis of the PHYDELIO randomized controlled trial[12] (EudraCT 2008-007237-47) — which originally assessed the efficacy of the cholinesterase inhibitor physostigmine for the prevention of postoperative delirium (POD) in patients undergoing major liver surgery—we investigated the perioperative dynamics of circulating autoantibodies (aABs) directed against five neurotransmitter-related G protein-coupled receptors (GPCRs): M3R, M4R, **β**_2_AR, D2R, and 5-HT_2_AR. Given the proposed role of neuroimmune dysregulation and neurotransmitter imbalance in the pathophysiology of POD, these specific aABs were selected for their relevance to cognitive function, autonomic regulation, and neuroinflammatory processes. The aim of this analysis was to determine whether major abdominal surgery elicits measurable changes in these GPCR aAB levels and whether such changes are associated with the incidence of POD or alterations in perioperative cognitive performance.

## Methods

### Study Design and Participants

Between August 11th, 2009, and March 3rd, 2016, 256 patients were recruited at Charité – Universitätsmedizin Berlin, Germany for the study “Perioperative Physostigmine Prophylaxis for Liver Resection Patients at Risk for Delirium and Postoperative Cognitive Dysfunction” (PHYDELIO, ISRCTN18978802; EudraCT 2008-007237-47) with 261 patients as a double-blind, randomized, placebo-controlled trial. [12] The 142-patient sample of the current explorative, cross-sectional comparison of antibody levels was a subsample of this larger project, including all patients for which the antibody levels were determined. Ethical approval was obtained by the ‘Landesamt für Gesundheit und Soziales Berlin’ (Berlin, Germany) ethics committee on 15 January 2009 (ZS EK 11 618/08), and written informed consent was obtained from all participants. Both the experimental results and the complete sample are described by Spies and colleagues.[12] Antibody level serum samples were available from all 142 patients at four time points: presurgically, as well as on postsurgical days 1, 2, and 7.

### Measures

#### Delirium

In line with Spies and colleagues [12] and in order to avoid underdiagnosis[13], we computed the cumulative delirium incidence (CDI) as our primary delirium endpoint, in which a patient is considered delirium positive, if they either had a) a positive Nursing Delirium Screening Scale score (NuDESC ≥ 2) in the recovery room or wards [14], b) a positive Confusion Assessment Method score for the ICU (CAM-ICU) [15], Delirium Detection Score (DDS ≥ 2) [14], or Intensive Care and Delirium Screening Checklist (ICDSC ≥ 4) score [16] in the ICU, c) a positive Delirium Rating Scale (DRS ≥ 18) [17], or Confusion Assessment Method score (CAM) [18] in the ward, or d) showed overall any signs of delirium (e.g. confused, agitated, drowsy, disorientated, delirious, received antipsychotic therapy) in the patient charts.

For descriptive purposes, we assessed presurgical cognitive impairments in line with the definition of Knaak and colleagues [19] and the ISPOCD criteria (see [12]).

#### Autoantibody levels in serum

Whole blood was collected in serum tubes, allowed to clot at room temperature for 45 minutes, and then centrifuged at 2500 × g for 10 minutes. The resulting serum was immediately stored at −80 °C. Circulating autoantibody levels were measured in the serum samples using a commercially available sandwich ELISA kit (CellTrend GmbH, Luckenwalde, Germany) according to the manufacturer’s instructions. Briefly, muscarinic acetylcholine receptors (M3R, M4R), **β**_2_AR, D2R, and 5-HT_2_AR. coated polystyrene plates were incubated with samples of a 1:100 serum dilution at 4°C for 2h. After washing, each sample was incubated with horseradish peroxidase-labeled goat anti-human IgG for 1h at room temperature. After washing, In the following enzymatic substrate reaction (room temperature, 20 min) the intensity of the colour correlates with the concentration of an anti-GPCR-antibody standard curve (2.5 to 40 U/ml).

### Statistical methods

We performed a retrospective analysis exploring the peri-surgical temporal course of autoantibody levels in serum and its effects on post operational delirium using a 2 *delirium* x 4 *time* x *antibody* mixed ANOVA approach, followed-up by a 2 *delirium* x 4 *time* mixed ANOVA analysis for each autoantibody receptor, separately. At each measurement-point, serum levels above two standard deviations from the mean were considered outliers and excluded from primary analyses (overall: n=25; M3R: n=12; M4R: n= 16; **β**2AR: n= 10; D2R: n= 8; 5-HT2AR: n= 13). Similar findings could be obtained for postsurgical M3R at day 1 and 2, comparing the delirium status with Wilcoxon-tests for each measurement instead of excluding outliers. Statistical tests were Greenhouse-Geisser corrected. The significance threshold was set at *p* < .05.

## RESULTS

### Sample characteristics

Overall, the 142 included patients were on average 58.9 years old (SD=11.7), 40.1% (57) of them were female, and 56.3% (80) were allocated to the experimental treatment condition. 11.3% (16) had identified presurgical cognitive impairments, and 45.8% (65) were postoperatively diagnosed with delirium according to CDI. This sample size is sufficiently powered (1-**β**) = .80 to find medium-sized effects of *d* ≥.47 with the set significance threshold.

### Temporal course of autoantibody levels

The course of autoantibody receptor serum levels was generally curvilinear across all measured antibodies (*time*: *F*(2.33,267.35) = 11.06, *p* < .001, **η**_part_^2^ = .088), albeit significantly differently pronounced across antibodies (*time x antibody*: *F*(3.70,425.04) = 7.15, *p* < .001, **η**_part_^2^ = .059): While M3R (*F*(2.45,313.93) = 7.57, *p* < .001, **η**_part_^2^ = .056), M4R (*F*(2.21,274.03) = 12.60, *p* < .001, **η**_part_^2^ = .092), D2R (*F*(2.37,313.17) = 15.54, *p* < .001, **η**_part_^2^ = .105), and 5-HT2AR (*F*(2.30,292.62) = 13.83, *p* < .001, **η**_part_^2^ = .098) significantly declined at the two measurement points after surgery and mostly recovered to presurgical levels on postsurgery day 7 (Figure 1), **β**2AR showed no significant change across time (*F*(2.55,331.12) = 2.01, *p* = .112, **η**_part_^2^ = .015).

**Figure 1.**
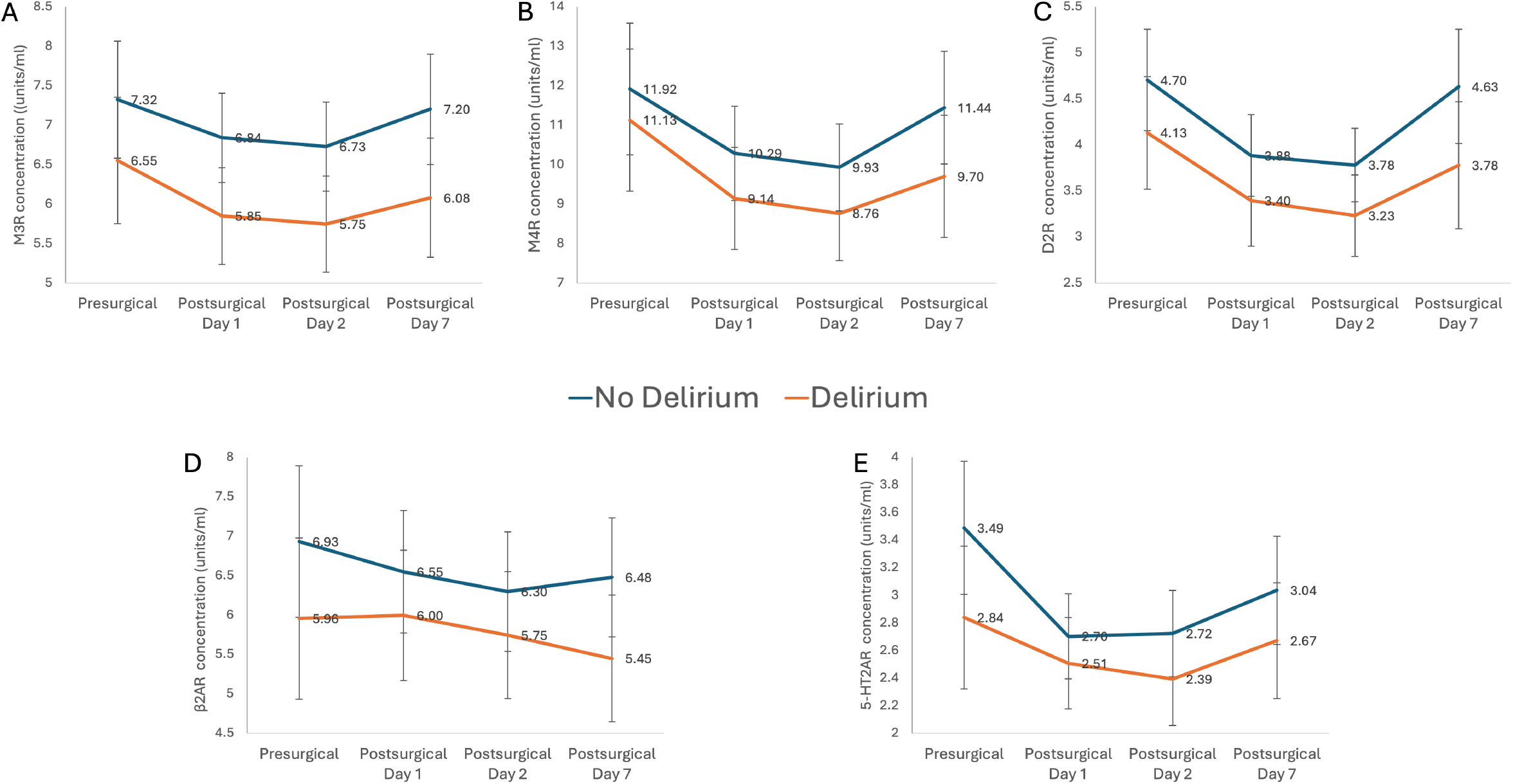
Peri-surgical course of autoantibody receptor serum level concentrations. *Notes*. Values denote mean levels and bars indicate 95% Confidence Interval

### Autoantibody levels and delirium

While delirium positive patients showed descriptively lower serum levels of all autoantibody receptors across all measurement points (Figure 1), the differences in these levels to the non-delirium patients reached significance solely for M3R antibodies (*F*(1,128) = 4.86, *p* = .029, **η**_part_^2^ = .037) and were neither significant across all antibodies (*F*(1,115) = 1.88, *p* = .174, **η**_part_^2^ = .016) nor for the remaining antibodies (all *F*≤ 3.39, all *p* ≥ .068,all **η**_part_^2^ ≤ .025).

This difference appeared to be temporally stable (overall: *F*(2.33,267.35) = 0.47, *p* = .655, **η**_part_^2^ = .004; M3R: *F*(2.45,313.93) = 0.40, *p* = .711, **η**_part_^2^ = .003; M4R: *F*(2.21,274.03) = 0.52, *p* =.613, **η**_part_^2^ = .004; **β**2AR *F*(2.55,331.12) = 0.68, *p* = .540, **η**_part_^2^ = .005; D2R *F*(2.37,313.17) = 0.53, *p* = .660, **η**_part_^2^ = .004, and 5-HT2AR *F*(2.30,292.62) = 1.64, *p* = .192, **η**_part_^2^ = .013). The experimental provision of physostigmine did not affect antibody receptor serum levels across all antibodies (*F*(3.70,425.04) = 7.15, *p* < .001, **η**_part_^2^ = .059) nor the temporal course of antibody receptor serum levels (*F*(3.70,425.04) = 7.15, *p* < .001, **η**_part_^2^ = .059).

## DISCUSSION

In this secondary analysis of the PHYDELIO randomized controlled trial, we investigated the perioperative dynamics of circulating autoantibodies (aABs) targeting five neurotransmitter-related GPCRs (M3R, M4R, **β**_2_AR, D2R, and 5-HT_2_AR) in patients undergoing major liver surgery. Our aim was to assess whether these aABs are modulated by surgical trauma and whether their levels are associated with the development of postoperative delirium (POD) or perioperative cognitive vulnerability.

Our findings demonstrate that major abdominal surgery leads to a transient decline in the serum levels of four out of five tested GPCR-specific aABs (M3R, M4R, D2R, and 5-HT_2_AR), with levels decreasing postoperatively and returning close to baseline by postoperative day seven. This curvilinear temporal pattern supports the notion that surgical trauma and the associated inflammatory response may transiently affect autoantibody production, receptor binding, or clearance mechanisms. It is conceivable that the peri-surgery procedure leads to dysfunction of organ barriers, influx of circulating autoantibodies and binding to GPCRs in the tissues, resulting in falling serum levels. A similar concept has been proposed for the brain acting as “immunoprecipitator” of autoantibodies after temporal dysfunction of the blood-brain barrier following cerebral ischemia or anesthesia.[20, 21] Interestingly, **β**_2_AR aAB levels remained stable throughout the perioperative period, suggesting receptor-specific regulatory dynamics or differential immunological relevance. Notably, antibodies against adrenergic and endothelin A receptors have previously been reported in patients with vascular and Alzheimer’s dementia.[22]

When examining the relationship between GPCR aAB levels and POD, we found that delirium-positive patients exhibited consistently lower levels of all measured autoantibodies. However, only M3R-specific aABs reached statistical significance in group comparisons. This finding aligns with the proposed role of muscarinic signaling dysfunction in the pathogenesis of POD and adds to the growing evidence implicating cholinergic modulation in perioperative neurocognitive disorders. Interestingly, while prior studies have reported elevated M3R aAB levels in various conditions, for instance in the context of Long Covid and cognitive dysfunction[23], a reduction in these antibodies in patients with delirium has not been previously described. Although speculative, M3R aAB may have beneficial or stabilizing effects for cognition under steady state conditions, while a drop in M3R aAB could then lead to cognitive dysfunction. Alternatively, M3R aAB serum levels may drop due to massive binding of the aABs to tissues after surgery. M3R aAB have been associated with acute postganglionic cholinergic dysautonomia, supporting their potential to disrupt cholinergic neurotransmission [24]. Additionally, M3R aABs have been implicated in secretory dysfunction in autoimmune diseases such as Sjögren’s syndrome, and have been linked to cognitive impairment and the pathogenesis of rheumatic diseases, including systemic sclerosis [25, 26].

Although only M3R-specific aABs reached statistical significance, the consistent directionality of effects across all GPCR aABs points toward a potential underlying signal that warrants further investigation. The effect sizes were small to moderate, which may have limited the ability to detect statistically significant associations for the other aABs.

Notably, the administration of physostigmine, a central cholinesterase inhibitor, did not alter aAB levels nor their temporal trajectories, indicating that modulation of cholinergic tone via enzymatic inhibition does not directly influence circulating GPCR aAB dynamics—at least within the examined timeframe and dosage.

## Limitations

This study has several limitations. First, the observed effects are exploratory and rely on a relatively liberal definition of postoperative delirium, which was not supported when more conservative definitions were applied. Second, as a secondary analysis, the study was not originally powered or designed to detect antibody-related effects, and the sample size—though sufficient for medium effect detection—may have limited sensitivity for small but biologically meaningful associations. Third, the selection of GPCR targets, while biologically plausible, was not exhaustive and excluded other potentially relevant receptors and signaling axes. Fourth, although ELISA-based assays provide quantitative data on aAB presence, they do not distinguish between agonistic and antagonistic functional activity, nor do they provide information about tissue distribution or receptor engagement in vivo.

In addition, while we accounted for several clinical covariates, we did not include comprehensive neuroimaging, cytokine profiling, or long-term cognitive follow-up, which could have provided more mechanistic insight and validated the clinical relevance of our immunological findings.

## Future Directions

Given the observed perioperative changes in autoantibody levels and the association between reduced M3R aAB concentrations and POD, future research should focus on elucidating the functional properties of these antibodies—particularly their agonistic or antagonistic effects—and how they interact with peripheral and central GPCR signaling pathways. Additionally, their potential role as biomarkers for POD risk stratification or as therapeutic targets in immunomodulatory strategies warrants systematic investigation. To this end, larger prospective studies incorporating multimodal biomarker profiling—such as neuroinflammatory cytokines, neuroimaging, and electrophysiological markers—alongside comprehensive cognitive assessments will be essential to determine whether GPCR autoantibodies are mechanistically involved in POD or serve primarily as peripheral indicators of broader neuroimmune dysregulation. Likewise, cloning and recombinant production of POD patient-derived monoclonal GPCR autoantibodies can provide valuable tools for the understanding of direct pathogenic effects (e.g. GPCR down-regulation, electrophysiological changes) *in vitro* and in animal models.

## Conclusion

This study provides initial evidence that major abdominal surgery transiently affects circulating GPCR autoantibody levels and that lower perioperative M3R aAB concentrations may be associated with the development of postoperative delirium. These findings support the growing view that neuroimmune dysregulation contributes to POD and highlight GPCR-directed autoantibodies as a potentially relevant, yet underexplored, factor in its pathogenesis. Further research is warranted to determine their functional role and diagnostic or therapeutic utility in perioperative neurocognitive disorders.

## Data Availability

All data produced in the present study are available upon reasonable request to the authors. Only anonymized data could be available.

## Acknowledgements

The authors sincerely thank Helge Giese, Cornelia Lachmann, and Klaus-Dieter Wernecke for their valuable support throughout the course and analysis of the PHYDELIO project.

